# Bone Morphogenetic Protein-9 Controls Pulmonary Vascular Growth and Remodeling

**DOI:** 10.1101/2023.06.02.23290910

**Authors:** Nihel Berrebeh, Yvon Mbouamboua, Raphaël Thuillet, Mina Ottaviani, Mustapha Kamel Chelgham, Virginie Magnone, Agnès Desroches-Castan, Nicolas Ricard, Ignacio Anegon, Séverine Remy, Ralph Theo Schermuly, Kevin Lebrigand, Baktybek Kojonazarov, Laurent Savale, Marc Humbert, Sabine Bailly, Pascal Barbry, Ly Tu, Christophe Guignabert

**Affiliations:** INSERM UMR_S 999 « Pulmonary Hypertension: Pathophysiology and Novel Therapies », Hôpital Marie Lannelongue, 92350 Le Plessis-Robinson, France; Université Paris-Saclay, Faculté de Médecine, Pulmonary Hypertension: Pathophysiology and Novel Therapies, 94276 Le Kremlin-Bicêtre, France; Université Côte d’Azur and CNRS, Institut de Pharmacologie Moléculaire et Cellulaire, F06560 Sophia Antipolis, France; Biosanté unit U1292, Grenoble Alpes University, INSERM, CEA, F-38000 Grenoble, France; Center for Research in Transplanation and Translational Immunology, Plateforme Trangénèse Rat & Immuno Phénomique, INSERM 1064 & SFR François Bonamy, CNRS UMS3556, F44098 Nantes, France; Department of Internal Medicine, Member of the German Center of Lung Research (DZL), Justus-Liebig-University of Giessen (JLU), Aulweg 130, Giessen, 35392, Germany; Assistance Publique - Hôpitaux de Paris (AP-HP), Service de Pneumologie et Soins Intensifs Respiratoires, Hôpital Bicêtre, 94270 Le Kremlin-Bicêtre, France

**Keywords:** pulmonary hypertension, pulmonary vascular remodeling, pulmonary vasodilation, angiogenesis, VEGF

## Abstract

**Background:** Pulmonary arterial hypertension (PAH), a life-limiting condition characterized by dysfunction of pulmonary microvascular endothelium, is predisposed by mutations in several genes that are critical for the proper activation of specific bone morphogenetic protein (BMP) receptor complexes that phosphorylate intracellular Smad1/5/8 in endothelial cells. However, the functional importance of BMP-9 (*GDF2*), one of the high affinity ligands for ALK1 (*ACVRL1*) and BMPR-II (*BMPR2*), for the pulmonary microvasculature remains imperfectly understood.

**Objective:** The aim of this study was first to determine the *in vivo* impact of BMP-9 deficiency on pulmonary vascular growth and remodeling, then to assess whether ALK1 expression can alter BMP-9 transcriptional signatures in human pulmonary microvascular endothelial cells (PMECs).

**Methods:** *CRISPR-Cas9* gene editing was used to create *Gdf2* knockout rats in *Sprague Dawley* background. Computed micro-tomography (Micro-Ct) scan after Microfil perfusion was performed to generate high-resolution 3D-images of the pulmonary arterial tree. The influence of ALK1 abundance on the transcriptional signatures of BMP-9 responses in human PMECs was assessed by single cell (sc)-RNAseq. Functional studies were performed using human PMECs exposed to BMP-9, the ALK1/2 inhibitor ML347, and ALK1-Fc fusion protein that neutralizes BMP9/10 and two animal models of severe pulmonary hypertension (PH).

**Results:** Micro-Ct angiography revealed structural and functional remodeling along the pulmonary vascular tree in BMP-9 deficient rats, resulting in vasodilation and increase in vascular density. scRNA-seq experiments identified distinct transcriptional signatures in human PMECs in response to BMP-9 responses. ALK1 expression had a direct impact on both proangiogenic capacities and transcriptional responses of PMECs to BMP-9. Functional studies performed in human PMECs confirmed that abundance of BMP-9 and ALK1 acted as modulators of PMEC tube formation, migration and proliferation, and also of vascular endothelial growth factor (VEGF)/VEGFR activities. The structural and functional remodeling observed in *Gdf2* knockout rats coincided with a lower susceptibility to develop severe PH induced by monocrotaline (MCT) and SU5416+hypoxia (SuHx).

**Conclusion:** BMP-9 and ALK1 are critical modulators of pulmonary vascular growth and remodeling. Our results provide potential mechanisms explaining why BMP-9 deficient animals are less susceptible to the rise in pulmonary vascular resistance in experimental models of PH.

## Non-standard Abbreviations and Acronym

α-smooth muscle actin: **α-SMA**; activin receptor-like kinase-1: **ALK1**; adrenomedullin: **ADM;** arbitrary unit: **AU**; protein kinase B: **AKT;** arteriovenous malformations: **AVMs**; bone morphogenetic protein: **BMP**; bone morphogenetic protein receptor type-II: **BMPR-II**; 5-bromo-2’deoxyuridine: **BrdU**; BMP response element: **BRE**; cluster: **C**; CRISPR-associated protein 9: **Cas9**; C-C motif chemokine ligand 2: **CCL2**; clusters of differentiation: **CD**; chronic hypoxia: **CHx**; clustered regularly interspaced short palindromic repeats: **CRISPR**; control: **CTR**; 4’,6-diamidino-2-phenylindole: **DAPI**; differentially expressed genes: **DEGs**; hereditary haemorrhagic telangiectasia: **HHT**; endothelial cell: **EC**; endothelin-1: **ET-1**; extracellular matrix: **ECM**; extracellular signal-regulated kinase 1/2: **ERK 1/2**, fetal calf serum, **FCS**; gene ontology/biological process: **GO/BP**; gene set enrichment analysis: **GSEA;** interleukin: **IL**; internal diameter: **ID**; knockout: **KO**; left ventricle: **LV**; low-density lipoprotein: **LDL**; lumen: **L**; messenger ribonucleic acid: **mRNA**; micro-computed tomography: **micro-CT**; monocrotaline: **MCT**; mean pulmonary artery pressure: **mPAP**; placental growth factor: **PGF**; pulmonary arterial hypertension: **PAH**; pulmonary artery-smooth muscle cell: **PA-SMC**; phosphatidylinositol 3-kinase: **PI3K**; pulmonary microvascular endothelial cell: **PMEC**; pulmonary vascular resistance: **PVR**; right ventricle: **RV**; single-cell RNA sequencing: **scRNA-seq**; sodium dodecyl-sulfate polyacrylamide gel electrophoresis: **SDS-PAGE**, sugen (SU5416)+chronic hypoxia: **SuHx**; vascular endothelial growth factor: **VEGF**; transforming growth factor-beta: **TGFβ**; von Willebrand factor: **vWF.**

## Clinical Perspective

**What Is New?**

- *Gdf2* knockout (KO) rats exhibit increased pulmonary vascular density and vasodilation compared to wild-type littermates.
- In experimental models of pulmonary hypertension (PH), *Gdf2* KO rats are less susceptible to the rise of pulmonary vascular resistance.
- BMP-9 has varying effects on different populations of human microvascular endothelial cells (PMECs), depending on the abundance of BMP receptors. Specifically, PMECs with higher levels of ALK1 exhibit a pro-angiogenic profile that is attenuated after BMP-9 stimulation.
- Our study demonstrated a complex interplay between BMP-9, ALK1, and vascular endothelial growth factor (VEGF)/VEGFR activities in PMECs.

**What Are the Clinical Implications?**

- BMP-9 deficiency alone may not be sufficient to induce spontaneous PH, underscoring the complex, multifactorial nature of the disease and highlighting the potential involvement of multiple hits, such as genetic and environmental factors. This underscores the need for a more comprehensive approach to understanding and treating PAH, including the use of molecules that target the interconnected BMP/TGFβ and VEGF/VEGR pathways.

## Introduction

Pulmonary arterial hypertension (PAH) is a severe cardiopulmonary vascular condition that can lead to right heart failure and death if left untreated. It is characterized by an increase in mean pulmonary arterial pressure (mPAP) at rest along with elevated pulmonary vascular resistance (PVR), resulting from marked pulmonary vascular remodeling ^1, 2^. This process involves narrowing and obliteration of pulmonary arterioles, accumulation of endothelial (ECs) and smooth muscle cells (PA-SMCs), and rarefaction of peripheral arterioles, leading to right ventricular (RV) pressure overload, dilatation, and failure. Despite current PAH-specific therapies, the 3-year survival rate remains at approximately 70% ^3^, highlighting the need for better treatments based on a deeper understanding of the disease pathophysiology.

Recent genetic studies have highlighted the crucial role of the loss of bone morphogenetic protein receptor type II (BMPR-II) signaling in the pathophysiology of human pulmonary arterial hypertension (PAH). In fact, loss-of-function mutations in genes encoding proteins that mediate BMPR-II signaling, such as *BMPR2*, *ACVRL1* (ALK1), *BMP10*, *ENG* (endoglin), *GDF2* (BMP-9), *CAV1* (caveolin-1), and *SMAD9* (Smad8), have been identified in heritable and sporadic cases of PAH ^4^. However, mutations in *ACVRL1*, *ENG*, or *SMAD4*, and occasionally in *GDF2*, are also associated with hereditary hemorrhagic telangiectasia (HHT), a congenital vascular patterning syndrome characterized by arteriovenous malformations (AVMs), mucocutaneous telangiectasia, and nosebleeds. Despite sharing common mutations affecting ligands, receptors, or signaling molecules of the BMPR-II pathway, HHT and PAH exhibit distinct phenotypes, penetrance, age of onset, and clinical management. To date, the underlying reasons for this dichotomy between these two conditions remain unclear.

The main objective of this study was to investigate the functional impact of BMP-9 deficiency, a high-affinity ligand for ALK1 and BMPR-II, on pulmonary vascular growth and remodeling *in vivo*, and to determine the significance of ALK1 abundance on the EC surface for the transcriptional response to BMP9 stimulation in human pulmonary microvascular endothelial cells (PMECs).

## Material and Methods

### Generation of *Gdf2* knockout (KO) rats

*Gdf*2-/-rats were generated using CRISPR-Cas9 technology by microinjection of a sgRNA complexed with Cas9 protein into *Sprague Dawley* rat zygotes ^5^. The targeted sequence for the sgRNA was on exon 2 (supplementary figure 1). Cleavage of the DNA generated a frameshift of the coding sequence with the generation of a premature stop codon defined by microfluidic capillary electrophoresis and Sanger DNA sequencing ^6^.

BRE (BMP Responsive element) dual luciferase activity assay was used to validate the deficiency in BMP-9 in cell lines. To this aim, NIH-3T3 cells were transfected with a mixture of the reporter plasmid pGL3(BRE)2-luc encoding firefly luciferase downstream of a BMP response element, pRL-TK luc encoding Renilla luciferase and a plasmid encoding human ALK1, as previously described ^7^. Rat sera (1% final) were added in serum free medium and luciferase measurements were performed 18 hours later.

### Assessment of pulmonary vascular changes

#### Contrast echocardiogram

Contrast-enhanced transthoracic echocardiography was blindly performed using microbubbles as ultrasound contrast agents generated by agitating a mixture of 9 mL Gelofusine^®^ solution (B, Braun) and 1 mL air between two 10 mL syringes connected by a 3-way stopcock. Once the microbubbles were prepared, 2 mL were injected as a bolus into the jugular vein that had previously been canulated with a fluid-filled polyethylene catheter for intravenous administration of agents. After a peripheral bolus injection, the passage of microbubbles through the right and left heart was visualized from a four-chamber apical view. To quantify the transit of microbubbles, we initially assessed the signal strength in decibels (dB) in the right ventricle before and during injection to ensure the uniformity of the administered microbubbles. Additionally, we measured the signal strength in dB in the left ventricle prior to injection and five heartbeats after the introduction of contrast microbubbles. We report the differences (Δ) observed in each ventricle before and after microbubble administration.

#### Microvessel perfusion

Fluorescent 8 µm microspheres (Phosphorex, Hopkinton, MA) were injected into the main pulmonary artery of anesthetized rats (isoflurane) to investigate right to left shunting (pulmonary arteriovenous shunts) as previously described ^8^. Then, we collected kidneys and brain from the injected rats and examined the trapped beads using Zeiss Axiovert A1 microscope (Zeiss, Paris, France). One kidney of each rat was minced, sonicated, and digested with proteinase K (0.2mg/mL) for 1 hour before the quantification of fluorescent microbeads using flow cytometry MACSQuant Analyser (Miltenyi Biotec, Paris, France).

#### Micro-computed tomography (micro-CT) imaging of the pulmonary artery tree

Rats were randomized, deeply anesthetized with isoflurane and heparinized. The pulmonary artery was canulated through the RV with a 21-gauge indwelling intravenous catheter filled with saline and the left atrium incised. To ensure maximum vessel dilation, the lungs were then perfused with 2 mL of sodium nitroprusside (10^-4^ M). The trachea was cannulated with PE-10 tubing and inflated to 20 cm H_2_O with 4% formaldehyde. Then, Microfil (Flowtech Inc, Carver, MA) was infused at 0.02 mL/min (8:1:1, solution of polymer: diluent: curing agent). After perfusion and solidification of the contrast medium, the trachea was ligated to maintain airway distension, and lungs were then removed *en bloc* and fixed in paraformaldehyde for 24 hours. An *ex vivo* imaging of the left lung was then performed by micro-CT system (Quantum GX, PerkinElmer, USA, Rigaku, Japan). Scan and analyses were performed blindly.

### Invasive right heart catheterization and echocardiographic assessment of left ventricular function

Rats were randomized and either studied in room air at 8-14 weeks of age, or after induction of mild PH upon exposure to chronic hypoxia (CHx). To validate our observations in models of severe PH, two different but complementary models were used: the monocrotaline (MCT) and sugen+hypoxia (SuHx) models. Briefly, *Gdf2*-/- and *Gdf2*+/+ rats were studied 3 weeks after a single subcutaneous injection of MCT (60 mg/kg; Sigma-Aldrich) or received a single subcutaneous injection of SU5416 (20 mg/kg) and were exposed to normobaric hypoxia for 3 weeks before returning to room air for 5 weeks. At the end of these protocols, pulsed-wave doppler during transthoracic echocardiography was used to evaluate the severity of PH by assessing pulmonary artery acceleration time (AT) to ejection time (ET) ratio and evaluate right ventricular function by assessment of the tricuspid annular plane systolic excursion (TAPSE) using Vivid E9 (GE Healthcare, Velizy-Villacoublay, France). In addition, mPAP were measured blindly by closed chest right heart catheterization, as previously described. A polyvinyl catheter was introduced into the right jugular vein and pushed through the RV into the pulmonary artery. In parallel, a carotid artery was cannulated for the measurement of systemic arterial pressure. Cardiac output (CO) in rats was measured using the thermodilution method. Hemodynamic values were automatically calculated by the physiological data acquisition system (LabChart 7 Software; ADInstruments Co., Shanghai, China). After measurement of hemodynamic parameters, the thorax was opened and the left lung immediately removed and frozen. The right lung was fixed in the distended state with formalin buffer. As previously described, the RV hypertrophy was assessed by the Fulton index [weight ratio of RV and (LV + septum)] and the percentage of wall thickness [(2 × medial wall thickness/ external diameter) × 100] and of muscularized vessels were determined ^9^.

#### Isolation, culture, and functional analysis of primary human pulmonary microvascular endothelial cells (PMECs)

Human PMECs were isolated and cultured as previously described ^10, 11^. Cells were routinely tested for mycoplasma and used at early passages < 5. To suppress BMP-9–ALK1–Smad1,5,8 signaling in human PMECs, studies were performed in the presence of human recombinant ALK1-Fc (R&D Systems), a ligand trap for BMP-9 and BMP-10, or with the ALK1 inhibitor ML347 (Tocris) at the concentrations indicated in the legends. Serum free medium or IgG1 in 5% FCS medium were used as controls.

#### siRNA transfection

To suppress ALK1 expression, human PMECs were transfected using lipofectamine RNAiMAX with 100 nM of ALK1 siRNA or with a scrambled sequence (Invitrogen, Cergy-Pontoise, France). The cells were studied within 3 days after transfection. Suppression of ALK1 levels was documented 72 hours after transfection.

#### Cell proliferation

EC proliferation was measured by 5-bromo-2′-deoxyuridine (BrdU) incorporation using the Delfia Cell proliferation kit (PerkinElmer, Courtaboeuf, France) and a time-resolved fluorometer EnVision Multilabel Reader (PerkinElmer) as previously described ^12^. Human PMECs were grown to 60% to 70% confluence at 37°C and synchronized for 24h in serum free medium, then exposed to recombinant BMP-9 (10ng/mL) (R&D Systems) in presence of 5% FCS for 24 hours.

#### Cell migration

EC migration was assessed using the *in vitro* wound-healing assay with the Ibidi Culture-Insert (Ibidi, Germany). Human PMECs were incubated with complete medium for 24h then after insert removal with or without BMP-9 10ng/mL in presence of 5% FCS.

#### Tube formation assay

Confluent monolayers of human PMECs were stained using Celltracker™ Green CFMDA (Invitrogen) for 30 min then exposed to 10 ng/mL of recombinant BMP-9 for 24h. Angiogenesis μ-slide (Ibidi, Germany) were coated with gel matrix (ECM from Engelbreth-Holm-Swarm murine sarcoma Sigma-Aldrich) 10µL/well. After 30 min polymerization at 37°C, suspension of cells (1×10^4^) pretreated with BMP-9 was added in a volume of 50 μL/well of medium with or without BMP-9. The cultures were left at 37°C for 4 hours and tube formation confirmed by fluorescent microscopy. Images were collected and parameters measured and quantified using Image J software.

### 10 X Single cell RNA Sequencing

#### Cell treatmen

Human PMECs from 5 explanted donor lungs were isolated, cultured (passage 3-4) and synchronized for 24 hours by serum starvation (0.3% FCS) and stimulated with 10 ng/ml BMP-9 for 4 hours, resulting in a total of 5 experimental conditions. Cell suspensions with viabilities between 96 and 98% were used for these experiments.

#### Sequencing

Single-cell profiling was performed with 10X Genomics Chromium kit RNA 3’ V3.1, according to the vendor specifications. Single-cell capture was performed according to a design of 5 experimental groups, each composed of one sample culture treated or not by BMP-9 from the same donor. Each condition in each experimental group was labelled with a distinct barcoded antibody, using the cell-hashing protocol developed by 10X Genomics. The five resulting single-cell libraries were sequenced as a paired-end run (R1:28bp x R2: 55bp) on an Illumina NextSeq500.

#### Single-cell data processing

10x Genomics libraries Fastq files were analyzed using Cell Ranger (v3.0.2) against hg38 genome. Hashtag oligonucleotides (HTOs) were counted using CITE-seq-Count v1.4.2^13^. Statistical analysis was performed using Seurat v4.0^14^. For each of the 5-sequencing run, cells from BMP9 stimulated and non-stimulated were demultiplexed based on their HTO enrichment using HTODemux Seurat function^15^ resulting in a total of 10 different samples (5 samples stimulated with BMP-9 and 5 samples non-stimulated with BMP-9). We first analysed each sample individually. Briefly, cells with less than 200 detected genes or more than 15% mitochondrial reads were removed. We normalized the data using SCTransform method (https://satijalab.org/seurat/articles/sctransform_vignette.html) and performed a dimensionality reduction by Principal Component Analysis (PCA) on the 3.000 highly variable features. Data were then visualized using Uniform Manifold Approximation and Projection (UMAP) embedding. Louvain algorithm^15^ and nearest neighbor graph, based on the first 30 principal components, was used to cluster the cells using a resolution of 0.3. For integration, we used reciprocal PCA method. Briefly, we separately integrated 5 samples stimulated with BMP-9 and 5 samples not stimulated with BMP-9. Next, we used the merge seurat function^14^ to group the two integrated datasets so that BMP-9 treated cells would not mix with untreated BMP-9 cells. We defined 4 clusters according to ACVRL1 (ALK1) expression: ALK1 high, ALK1 high + BMP-9, ALK1 low and ALK1 low + BMP-9. Identification of gene markers was done using FindAllMarkers Seurat function^14^. Differential analysis between clusters were carried out with FindMarkers Seurat function^14^. Functional analysis and Gene Set Enrichment Analysis was then done using gene markers using ClusterProfiler package and gseGO function^16^.

### Total RNA Isolation and Real-Time Quantitative Polymerase Chain Reaction

The mRNA expression of *ACVRL1* were measured by real-time quantitative PCR as previously described ^11^.

### Western immunoblot and Immunostaining

Cells/tissues were homogenized and sonicated in RIPA buffer containing protease and phosphatase inhibitors (Sigma-Aldrich). Protein extract (30 µg) was used to detect ALK1, VEGFR1, VEGFR2, p-VEGFR2, p-SRC, p-AKT, AKT, GAPDH and β-actin (**Table S1**) by SDS-PAGE as previously described ^11^. For immunohistochemistry and immunofluorescent staining, paraffin sections were incubated overnight at 4°C with antibodies directed against ALK1, alpha-smooth muscle actin (α-SMA), von Willebrand factor (vWF), CD31 (**Table S1**) as previously described ^11^.

### Statistical Analyses

All results are presented as mean ± SEM. Statistical calculations were performed with GraphPad Prism 7 (GraphPad Software, Inc). The unpaired Student t test was used after testing for normality and equal variance (Shapiro-Wilk test) to compare 2 groups. One-way ANOVA Tukey post hoc test was used to compare multiple groups if the data followed a normal distribution, otherwise nonparametric Kruskal-Wallis post hoc tests were used. Sample size is indicated in the figure legends. Results with *P*<0.05 were considered statistically significant.

### Ethics statement

Animal experiments were approved by the Ethics Committee of the Université Paris-Saclay and carried out in accordance with the Guide for the Care and Use of Laboratory Animals adopted by the National Institute of Health and Medical Research (Inserm) and were registered with ministerial numbers APAFiS #35319. The use of human tissues was approved by the local ethics committee (CPP EST-III n°18.06.06, Le Kremlin-Bicêtre, France) and all patients gave informed consent before the study.

## Results

### Assessment of the pulmonary arterial bed in BMP-9 deficient rats

We found that *Gdf2*-/-rats were viable and fertile and developed normally with no obvious phenotypic abnormality. To visualize and determine whether BMP-9 deficiency is associated with defects in the pulmonary arterial bed in *Gdf2*-/-KO rats, *Gdf2-/-* and *Gdf2*+/+ rats were terminally perfused with Microfil via the main pulmonary artery and left lung were subjected to micro-Ct imaging and analysis. The loss of BMP-9 signaling was validated by the loss of BRE activity (**Figure S1**). We found higher vessel volume and numbers of vascular junctions in *Gdf2-/-* rat lungs relative to *Gdf2*+/+ rat lungs (**Figure 1A**). These vascular differences were associated with decreased levels of hemoglobin and hematocrit in *Gdf2-/-* relative to *Gdf2*+/+ rats (**Figure 1B**). To further characterize this vascular phenotype in BMP-9 deficient rats, we injected a microbubble-based contrast agent in the pulmonary artery to perform ultrasound imaging technique. Figure 1C shows two-D echocardiography in apical four chamber view demonstrating microbubbles in the right-sided chambers of the heart in both *Gdf2*-/- and *Gdf2*+/+ rats. However, microbubbles were also observed in the left chamber of the heart in *Gdf2*-/-rats originating from the pulmonary vein 5 heart beats following injection (**Figure 1C**), indicating vasodilation and/or the existence of arteriovenous shunting in the pulmonary vasculature. We confirmed these observations with the passage of fluorescent microbeads (8 µm in diameter) from the jugular vein to the systemic vasculature, as reflected by the higher numbers of microbeads per kidney found in *Gdf2*-/-rats relative to *Gdf2*+/+ rats (**Figure S2**). Consistent with these vascular differences, higher numbers of small-diameter (internal diameter [ID] < 20 µm) and large-diameter vessels (20 µm < ID < 200 µm) were found (**Figure 1D**).

**Figure 1:**
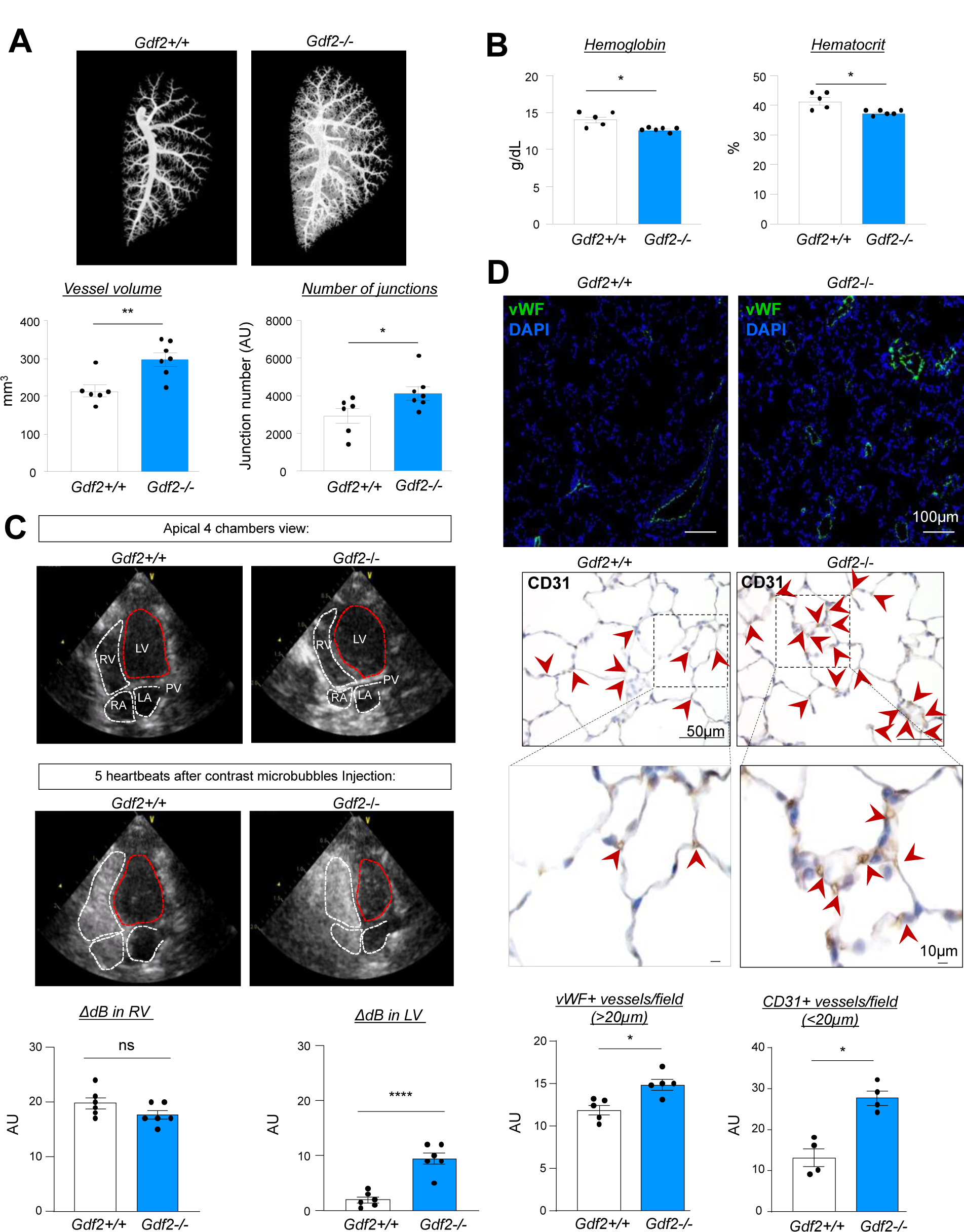
*Gdf2-/-* rats exhibit structural and functional abnormalities of the pulmonary vascular bed. **(A)** representative images using X ray micro-angiography showing the left lung pulmonary vascular bed with quantification of the vessel volume and the number of junctions. **(B)** Hemoglobin and hematocrit measurements showing a decrease in *Gdf2*-/-rats. (**C)** contrast enhanced ultrasound imaging and quantification of the passage of microbubbles in the left ventricle in *Gdf2-/-* rats. **(D)** Quantification of pulmonary vessel density based on CD31 and von Willebrand factor (vWF) staining expressed as the number of positive vessels measured in 20 fields. Red arrows represent CD31+ cells. Data are represented as mean± SEM. Comparisons were made using the nonparametric Mann-Whitney t-test. * p<0.05; **, p<0.01; ****, p<0.0001. Scale bar = 50µm for CD31 staining and 100µm for vWF immunostaining. AU = arbitrary unit. DAPI = 4’,6-diamidino-2-phenylindole. ΔdB = change in decibel levels. LV = left ventricle. RV = right ventricle. ns = non significant.

These data thus reveal that BMP-9 deficiency in rats led to structural and functional remodeling along the pulmonary vascular tree, resulting in vasodilation and increase in density.

### Determination of the transcriptomic responses of human pulmonary microvascular EC (PMEC) subtypes to BMP-9

To address whether distinct transcriptional signatures of BMP-9 responses are present among the different types of pulmonary ECs and if a relationship exist with the abundance of type-1, type-2, and type-3 receptors, primary cultures of human PMECs were generated from lung specimens. The BMP-9 transcriptional responses were analyzed at early passages (< 4) in 5 healthy donors (n=5) using scRNA-seq. At baseline, these human cells represent 5 distinct PMEC populations that naturally expressed different patterns of BMP/TGF-β receptors that can therefore be used to study how they specifically respond to exogenous BMP-9 (**Figure 2A**). In the absence of BMP-9, these 5 distinct clusters indeed expressed different levels of ALK1 (*ACVRL1*), BMPR-II (*BMPR2*), TGFβRII (*TGFBR2*), and endoglin (*ENG*). However, C5 (referred as ALK1^high^) expressed high levels of ALK1 (*ACVRL1*) and BMPR-II (*BMPR2*), which bind with strong affinity BMP-9, compared to C1-C4 (referred as ALK1^low^) (**Figure 2A**).

**Figure 2:**
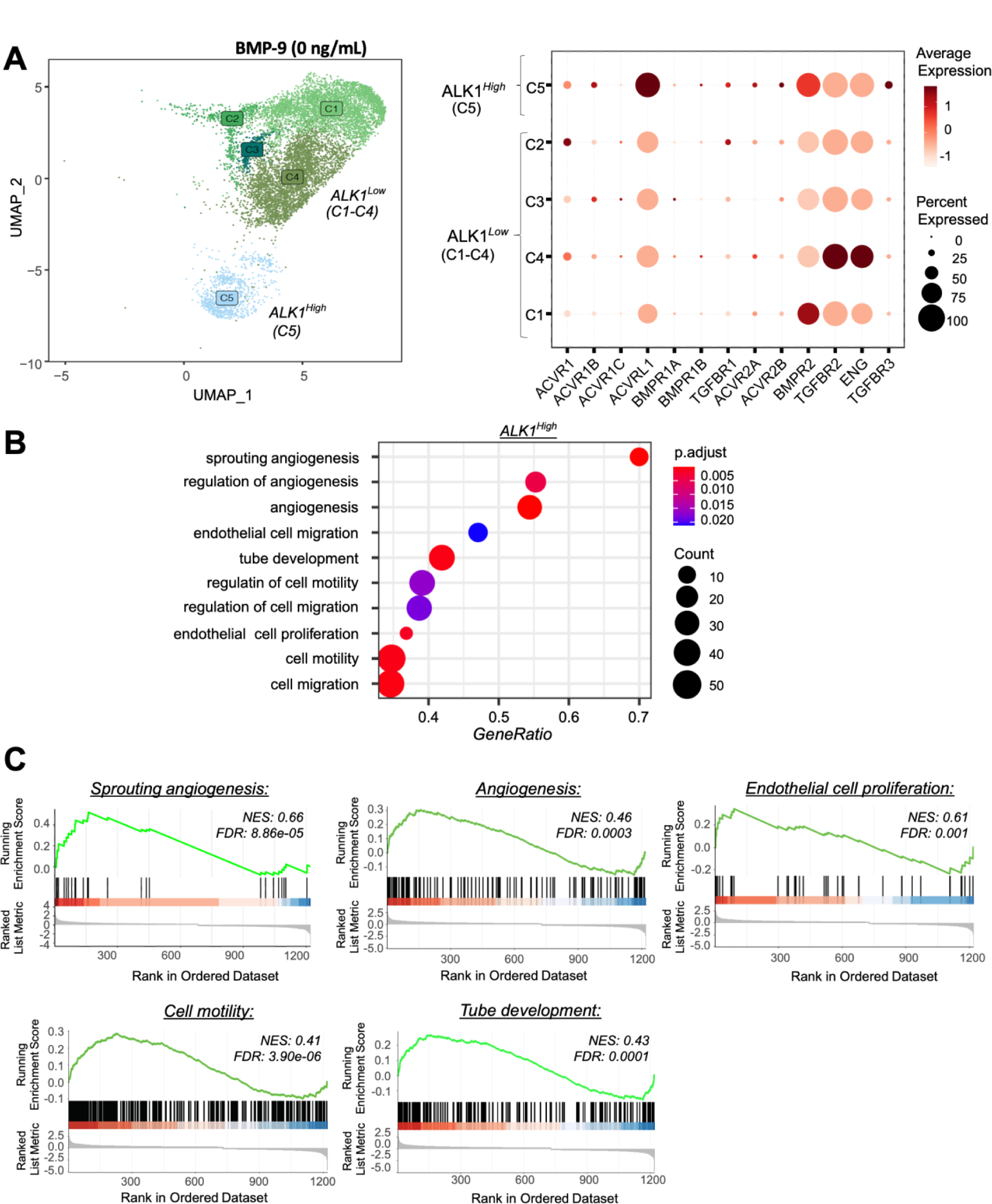
scRNA-seq analyses revealed 5 distinct populations of human pulmonary microvascular ECs (PMECs) that naturally expressed different patterns of BMP/TGF-β receptors in primary EC cultures derived from human lung tissue. (**A**) Transcriptomic profile of each cell (dots) represented by a Uniform Manifold Approximation and Projection (UMAP), noted from C1 to 5 in the basal state and differential expression of BMP receptors in the five clusters (n=5), the clusters can be subdivided on two subgroups based on ALK1 expression: ALK1 ^high^ and ALK1 ^low^. **(B)** TOP 10 GO terms that characterized the ALK1^high^ PMEC population. **(C)** Gene set enrichment analysis (GSEA) of differentially expressed genes (DEGs) between ALK1^high^ *versus* ALK1^low^ PMECs. NES: normalized enrichment score; FDR: false discovery rate.

To better characterize the cellular phenotype in ALK1^high^ and ALK1^low^ PMEC-clusters, we compared gene ontology term enrichment between the two clusters using the functional *Gene Ontology/Biological Process* (GO/BP) database. The ALK1^high^ PMEC-cluster was characterized by a strong proangiogenic signature, as reflected by the expression of genes involved in the activation of critical biological processes and molecular pathways, such as sprouting angiogenesis, regulation of angiogenesis, cell motility, tube development, and EC proliferation and migration (**Figure 2B and S3A**). A similar observation was made after a gene set enrichment analysis (GSEA) of differentially expressed genes (DEGs) between ALK1^high^ PMECs *versus* ALK1^low^ PMECs showed that also identified sprouting angiogenesis, angiogenesis, EC proliferation, cell motility and tube development (**Figure 2C**). We found several genes belonging to the VEGF pathway to be highly expressed in this ALK1^high^ PMEC-cluster, such as VEGFR2 (*KDR*), placental growth factor (*PGF*), VEGF B, VEGF C, and the decoy receptor VEGFR1 (*FLT1*) (**Table S2**). In contrast, the TOP 50 GO terms in ALK1^low^ PMEC-clusters contained terms associated with negative regulation of fibroblast proliferation, fatty acid transport, triglyceride metabolic process, lipid transport, tissue morphogenesis circulatory system development, suggesting that ALK1^low^ PMECs are most likely involved in vascular stability (**Figure S3B**). Of note, all PMEC-clusters identified displayed comparable levels of different EC markers, including of CD31 (*PECAM1*), ETS-related gene (*ERG*), cadherin-5 (*CDH5*), or Tie2 (*TEK*) (**Figure S4A**) and were negative for mesenchymal markers such as SM22 (*TAGLN*) or α-smooth muscle actin (*ACTA2*) (**Figure S4B**).

A 4-hour exposure to 10 ng/mL of recombinant BMP-9 markedly altered the transcriptome of each PMEC-clusters with no change in their respective identity, as revealed by the identification of distinct clusters of cells exposed to BMP-9 (**Figure 3A and S3**). We observed that BMP-9 treatment resulted in a decrease in *ACVRL1* mRNA levels, as well as an expected increase in the expression of several BMP-9 target genes in both ALK1^low^ and ALK1^high^ PMECs. Specifically, we found increased expression of mRNA for *BMPR2*, *ENG*, *ID1*, *ID2, ID3*, *SMAD6*, and *SMAD7* in response to BMP-9 treatment (**Figure 3B, S4C and Table S2**). Furthermore, the exposure of ALK1^high^ PMEC-cluster to BMP-9 leads to attenuation of the proangiogenic signature as reflected by the loss of pathways/gene sets related to angiogenesis in the GO/BP analysis (**Figure 3C**). The GO/BP analysis between ALK1^low^ + BMP9 *versus* ALK1^low^ revealed that changes in gene expression induced by BMP-9 in ALK1^low^ are associated with regulation of BMP signaling pathway, regulation of blood pressure, regulation of ossification, endothelium development, or blood vessel development (**Figure 3D**). Of note, each cluster preserved its endothelial identity and displayed similar mRNA levels of *PECAM1*, *ERG*, *CDH5*, *TEK, SNAI1, SNAI2, or TWIST1* markers, without mesenchymal marker expression (**Figure S4**).

**Figure 3:**
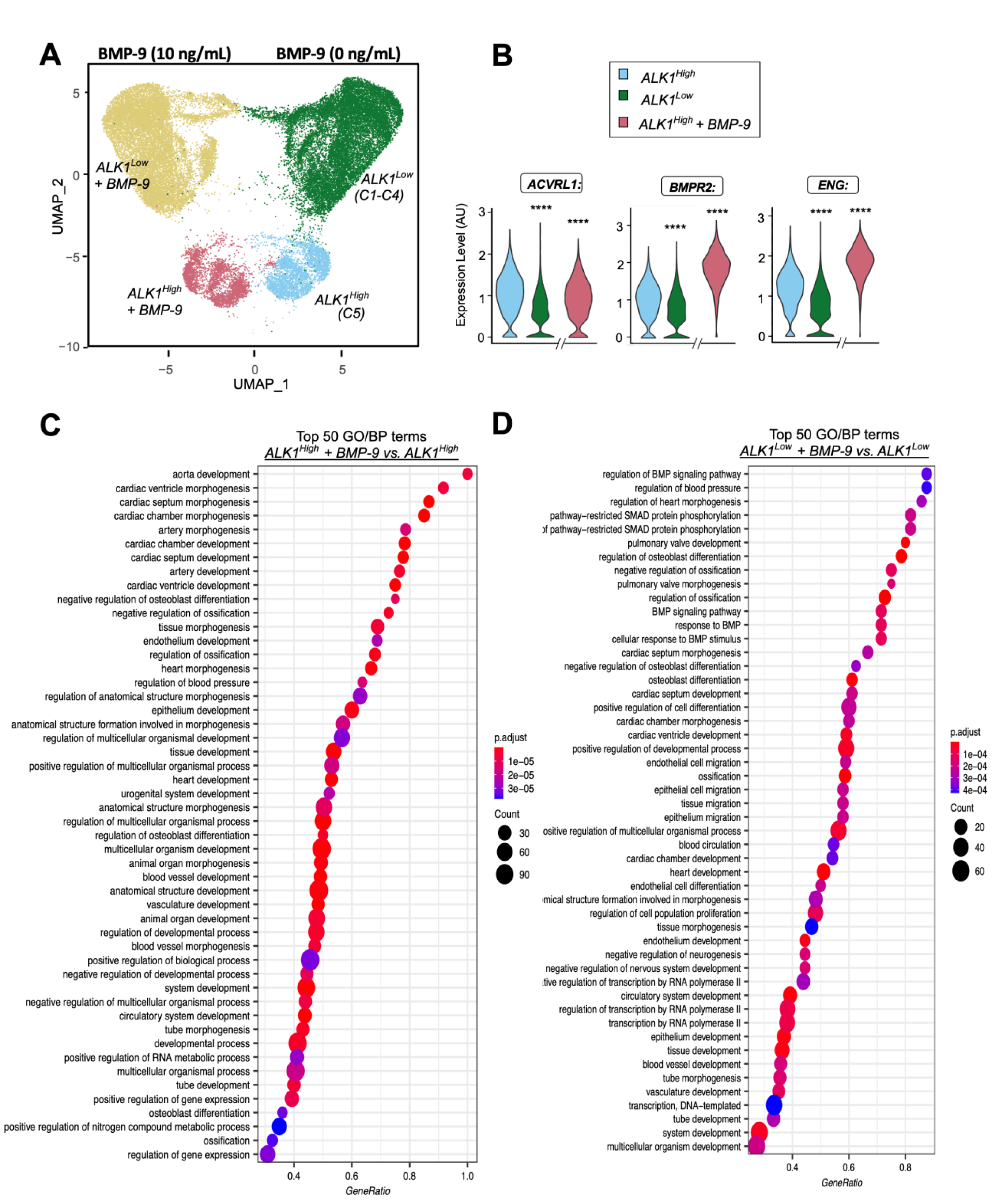
Presence of distinct transcriptional signatures of BMP-9 responses in cultured human pulmonary microvascular ECs (PMECs). **(A)** UMAP illustration showing changes in the transcriptomic profile in ALK1^high^ and ALK1^low^ PMEC-clusters 4h after BMP-9 stimulation (10ng/mL). **(B)** Violin plots of standardized expression of the main BMP-9 receptors across ALK1^high^ and ALK1^low^ PMEC-clusters with or without BMP-9 stimulation. TOP 50 *Gene Ontology/Biological Process* (GO/BP) terms that characterized the BMP-9 responses in ALK1^high^ **(C)** and ALK1^high^ PMEC-clusters (**D)**. Data are represented as mean± SEM. Significance was measured using nonparametric Mann-Whitney t-test: ****, p<0.0001 compared to ALK1^high^.

The scRNA-seq data obtained in cultured human PMECs indicate the existence of distinct transcriptional signatures of BMP-9 responses in pulmonary microvascular ECs which are supported, at least in part, by ALK1 abundance that both affects the EC phenotype and BMP-9 transcriptional responses.

### Role of ALK1 as a modulator of PMEC tube formation, migration and proliferation

To investigate the role of ALK1 as a modulator of PMEC tube formation, migration, and proliferation, we examined the effects of decreasing BMP-9 and ALK1 levels using ALK1-Fc and siRNA, respectively, on cultured human PMECs. Our results showed that the addition of exogenous BMP-9 attenuated cell migration, proliferation, and tube formation, consistent with previous studies ^17^ (**Figure S5**). Consistent with these findings, removing BMP-9 from media containing 5% FCS using ALK1-Fc showed the opposite effects on PMECs (**Figure 4A-C**). We also confirmed that decreasing the abundance of ALK1 in serum enriched media could reduce PMEC migration, proliferation, and tube formation.

**Figure 4:**
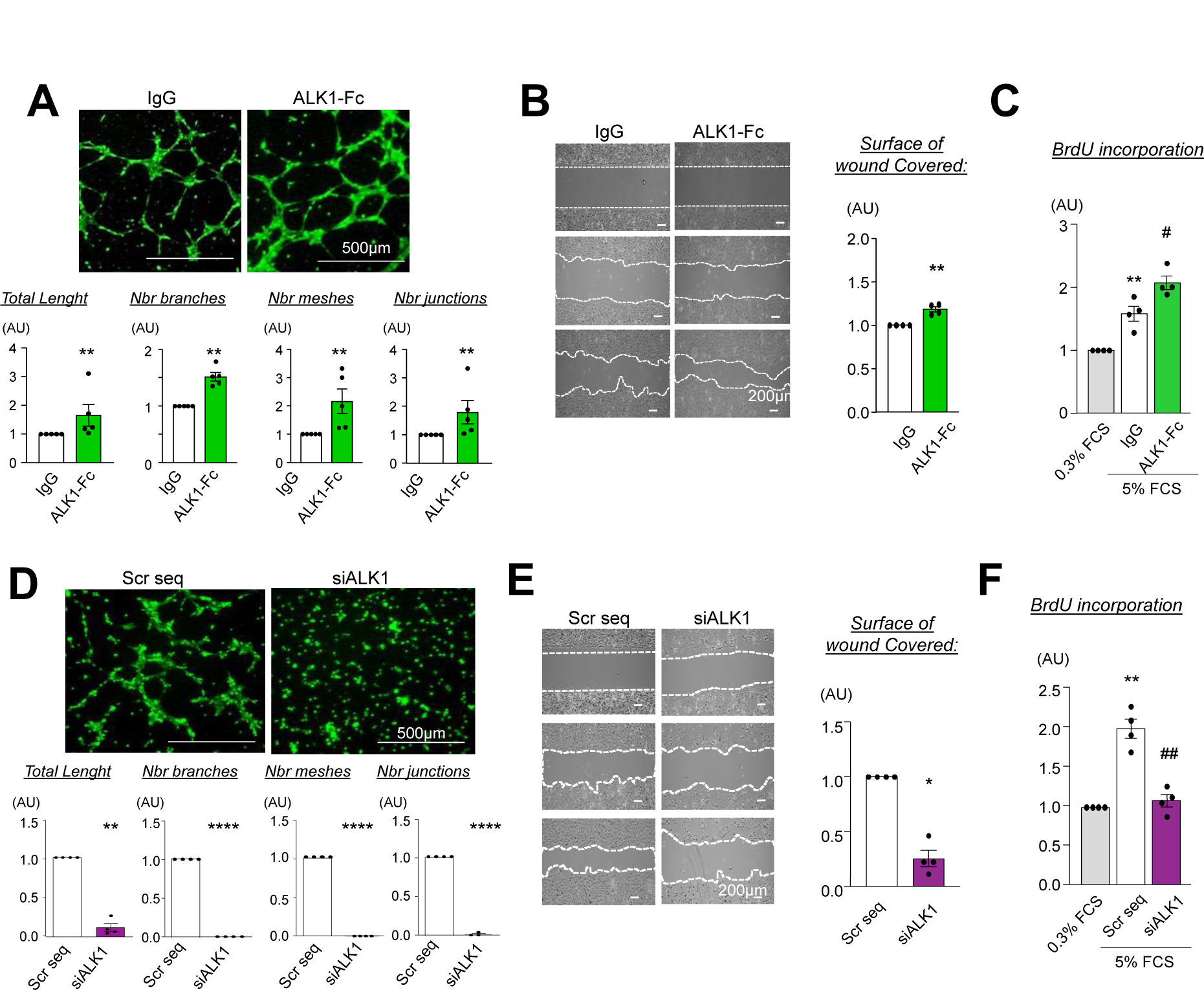
Impact of decreasing BMP-9 and ALK1 levels using ALK1-Fc and siRNA on the capacity of human PMECs to form tubes and to their migratory and proliferative potentials. **(A)** Representative images and quantifications of tube formation by human PMECs exposed to non-relevant IgG or ALK1-Fc (300ng/mL). **(B)** Representative images and quantifications of the surface of wound covered by human PMECs exposed to non-relevant IgG or ALK1-Fc (300ng/mL). **(C)** Effects of BMP-9 and siALK1 on BrdU incorporation in human PMECs. **(D-F)** Effects of non-relevant IgG or ALK1-Fc (300ng/mL) on tube formation, migration and proliferation of human PMECs. Data are represented as mean± SEM. Significance was measured using parametric paired t-test or 1-way ANOVA with Tukey post hoc tests: *, p<0.05; **, p<0.01; ****, p<0.0001 *versus* IgG, Scr sequence or 0.3% fetal calf serum (FCS). #, p<0.05; ##, p<0.01 *versus* Scr seq. AU: arbitrary unit. ALK1-Fc: soluble chimeric protein consisting of the extracellular part of ALK1 fused to a Fc fragment. IgG: immunoglobulin G. Nbr: number. BrdU: bromodeoxyuridine

Specifically, siRNA-mediated knockdown of ALK1 (**Figure 4D-F and Figure S6**) and inhibition with the potent and selective ALK2 and ALK1 inhibitor ML 347 (**Figure S5**) both resulted in reduced PMEC migration, proliferation, and tube formation. These findings indicate that, similar to BMP-9 levels, the abundance of ALK1 plays a significant role in PMEC tube formation, migration, and proliferation.

### In vitro and in vivo modulation of VEGF/VEGFR activities by BMP-9 and ALK1

Our scRNA-seq data reveals that PMECs with lower expression of ALK1 (ALK1^low^) have decreased expression of FLT1 and KDR receptors, as well as lower production of PGF, compared to PMECs with high ALK1 expression (ALK1^high^). In addition, we found that treatment of ALK1^high^ PMECs with BMP-9 results in increased expression of VEGF family ligands, such as VEGF-C and PGF, and strongly induces the expression of the FLT1 receptor (**Figure 5A**). To confirm the link between BMP-9 and the VEGF signaling pathway, we conducted Western blot analysis to assess the effects of BMP-9 on ALK1, VEGFR1, and VEGFR2 protein levels. Our results showed that BMP-9 decreases ALK1 protein levels in human PMECs (**Figure 5B**), while simultaneously increasing VEGFR1 (∼2-3 fold) and VEGFR2 (∼1.5-2 fold) expression (**Figure 5C**). Furthermore, BMP-9 pretreatment of human PMECs attenuates VEGFR2, Src, and Akt phosphorylation in human PMECs exposed to VEGF (**Figure 5D**). Our findings are consistent with the observation that *Gdf2*-/-rat lungs display higher ALK1 protein levels and VEGF/VEGFR activities relative to *Gdf2*+/+ (**Figure 5E and 5F**), which suggests that the abundance of ALK1, like BMP-9 levels, is an important modulator of VEGF/VEGFR activities in PMECs both *in vitro* and *in vivo*.

**Figure 5:**
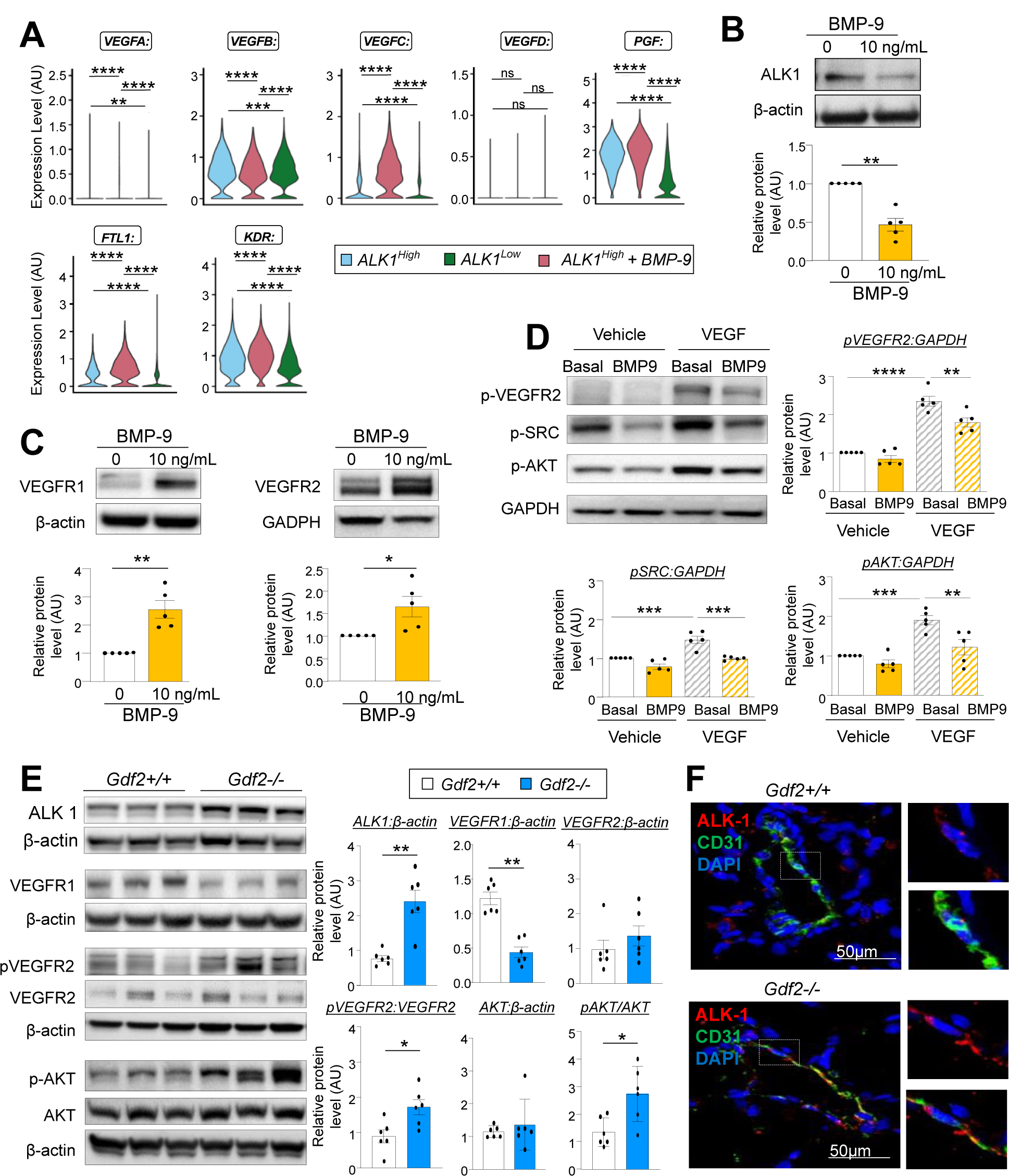
BMP-9/ALK1 modulation interferes with the VEGF signaling system. **(A)** Violin plots showing BMP-9 effects on mRNA levels for VEGF-A, VEGF-B, VEGF-C, VEGF-D, placental growth factor (*PGF*), VEGFR1 (*FLT1*), and VEGFR2 (*KDR*) in ALK1^High^, with or without BMP-9 stimulation and in ALK1^Low^. Representative immunoblots and densitometric analysis of ALK1**(B)**, VEGFR2 and VEGFR1 expression in human PMECs upon 24h of BMP-9 stimulation (10ng/mL) **(C)**. **(D)** Representative immunoblots and densitometric analysis of p-VEGFR2, p-SRC and p-AKT in human PMECs pretreated for 24h with BMP-9 (10ng/mL) and/or stimulated for 30min with VEGF-A (20ng/mL). **(E)** Representative immunoblots and densitometric analysis of ALK1, VEGFR1, p-VEGFR2, VEGFR2, p-AKT and AKT in *Gdf2-/-* and *Gdf2+/+* rat lungs. **(F)** Representative immunofluorescent staining of ALK1 (red), CD31 (green) and nuclei (DAPI, blue) in *Gdf2-/-* and *Gdf2+/+* rat lungs. Scale bars=50 μm. Data are represented as mean± SEM. Significance was measured using parametric paired t-test or 1-way ANOVA with Tukey post hoc tests: *, p<0.05; **, p<0.01; ***, p<0.001; ****, p<0.0001 *versus* basal condition or *Gdf2*+/+. AKT = protein kinase B. AU = arbitrary unit. DAPI = 4’,6-diamidino-2-phenylindole. GAPDH = glyceraldehyde-3-phosphate dehydrogenase

### *Susceptibility of* BMP-9 deficient rats to pulmonary vascular remodeling induced by monocrotaline (MCT) and sugen+hypoxia (SuHx)

*Gdf2*-/-rats exhibit normal values of mPAP as compared with *Gdf2*+/+ rats (15.3 ± 0.4 *versus* 15.7 ± 0.4 mmHg, respectively, NS), cardiac output (94.5 ± 1.8 *versus* 100.3 ±1.6 mL/min, respectively, NS), and total pulmonary vascular resistance (161 ± 5 *versus* 156 ± 4 mmHg/mL/min, respectively, NS) (**Figure 6**). Consistent with these findings, no obvious changes were observed in blood vascular structures in lungs of *Gdf2*-/-rats relative to *Gdf2*+/+ rats (% of wall thickness: 23 ± 1.7 versus 23 ± 1.6 %, respectively, NS; % of muscularized arteries: 11.2 ± 0.9 *versus* 11.9 ± 0.9 %, respectively, NS) (**Figure 7A**). In addition, no difference in cardiomyocyte size (146 ± 36 *versus* 163 ± 8 µm^2^, respectively, NS) and in collagen deposition were noted between *Gdf2*-/- and *Gdf2*+/+ rats using picrosirius staining (2.5 ± 0.4 *versus* 1.9 ± 0.4 %, respectively, NS) (**Figure 7B**). Consistent with previously published data obtained in BMP-9 deficient mice chronically exposed to hypoxia ^18, 19^, we also found that *Gdf2*-/-rats developed less pronounced pulmonary vascular remodeling and PH induced by chronic hypoxia than *Gdf2*+/+ rats (**Figure 6 and 7**).

**Figure 6:**
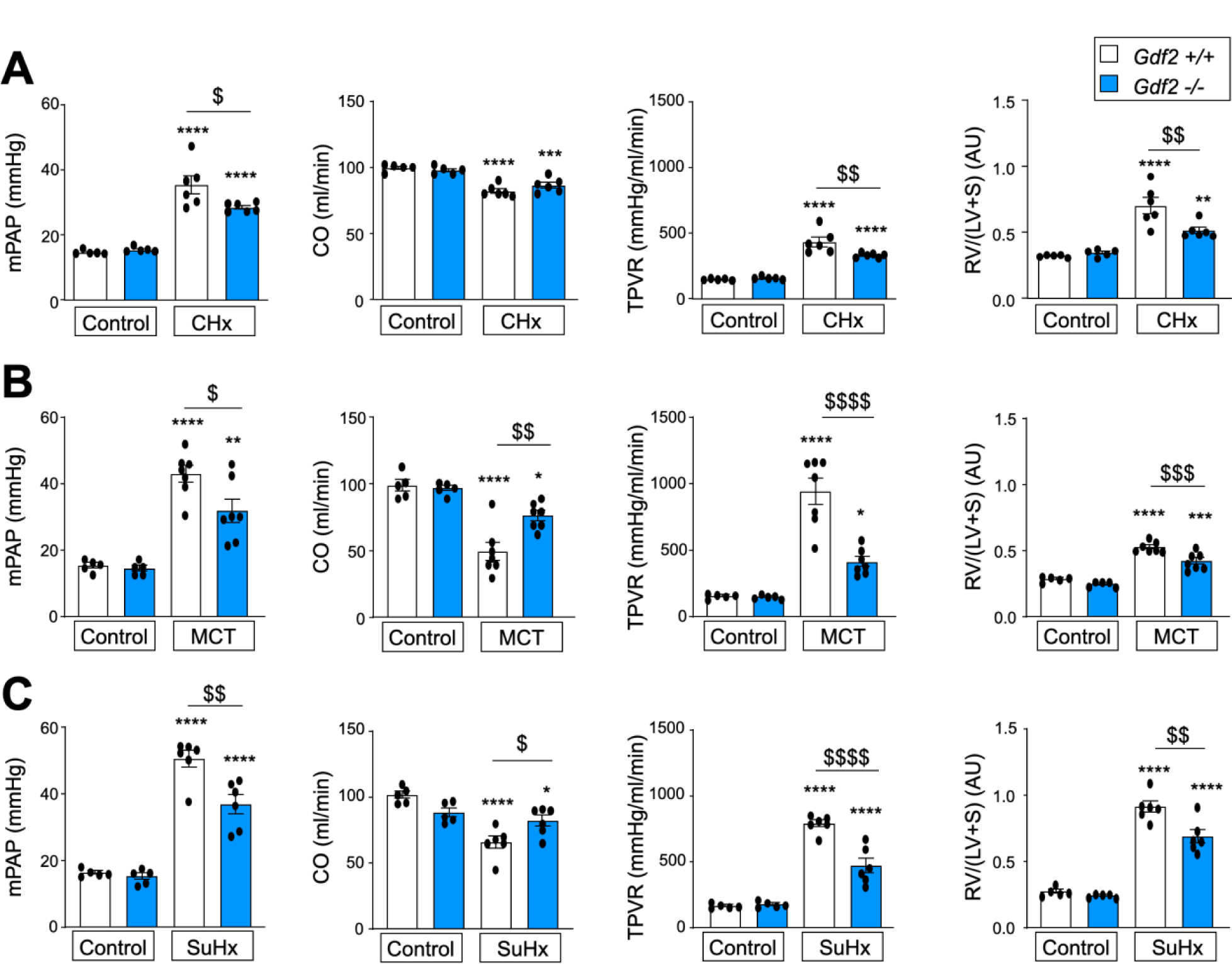
*Gdf2-/-* rats show less severe pulmonary hemodynamic profile as compared to *Gdf2+/+* in experimental models of pulmonary hypertension (PH). Mean pulmonary arterial pressures (mPAP), total pulmonary vascular resistance (TPVR), cardiac output (CO), and right ventricular hypertrophy (evaluated by the Fulton index) **(A)** in the chronic hypoxia, **(B)** the monocrotaline (MCT), and **(C)** the Sugen+hypoxia (SuHx) rat models of severe PH. Significance was measured using 1-way ANOVA with Tukey post hoc tests. *, p<0.05; **, p<0.01; ***, p<0.001; ****, p<0.0001 *versus* control *Gdf2+/+* rats $, p<0.05; $$, p<0.01; $$$, p<0.001; $$$$, p<0.0001 *versus Gdf2*+/+ CHx, MCT, or SuHx rats.

**Figure 7:**
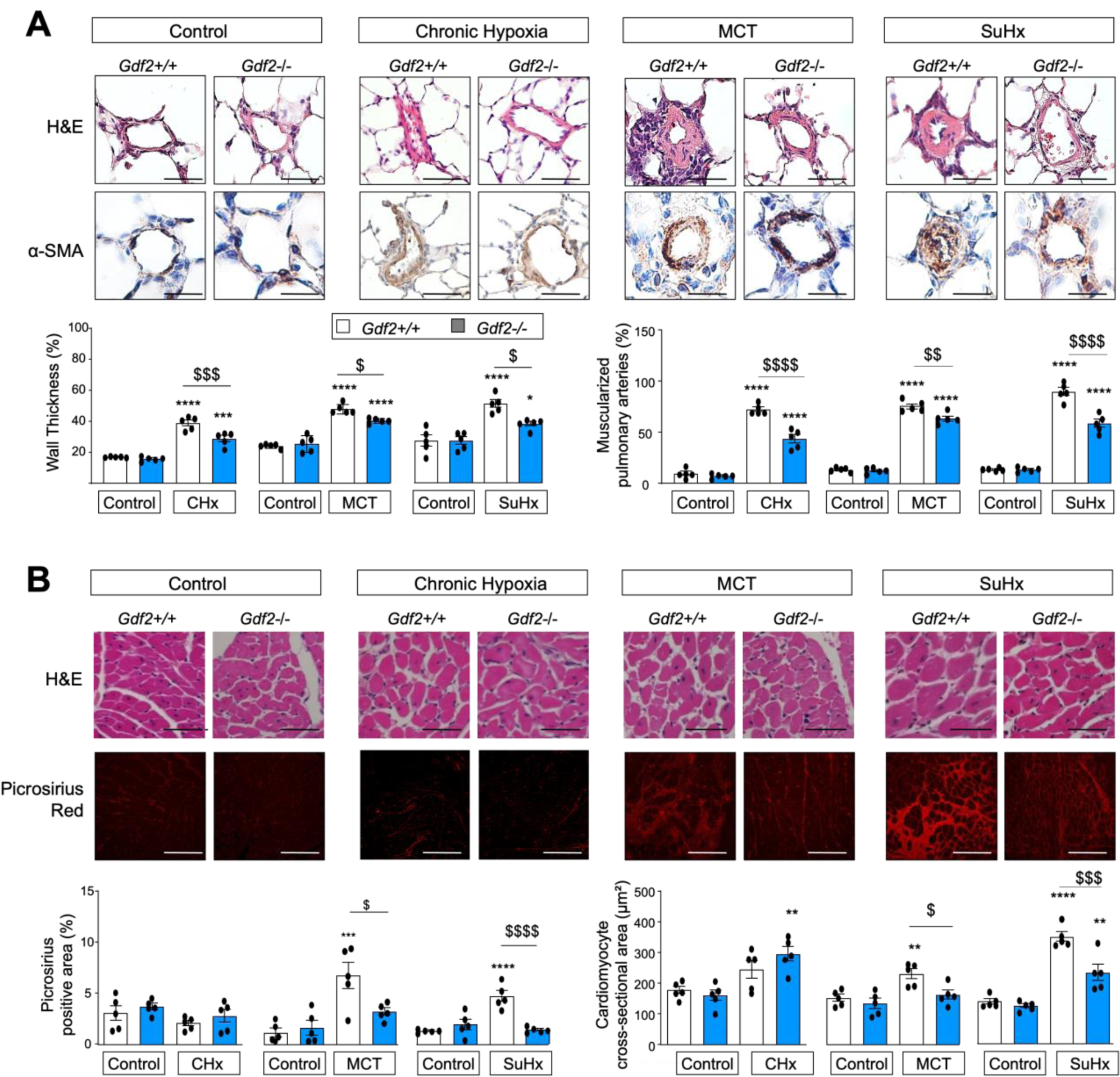
*Gdf2-/-* rats exhibit less pronounced cardiac and pulmonary vascular remodeling as compared to *Gdf2+/+* experimental models of pulmonary hypertension (PH). **(A)** Representation of hematoxylin and eosin (H&E), alpha-smooth muscle actin (α-SMA) staining, and quantification of the percentage of wall thickness and of muscularized pulmonary arteries in lungs from *Gdf2*-/- and *Gdf2*+/+ rats. (**B)** Representative images of H&E and picrosirius staining of tissue sections of right ventricle myocardium from either control, chronic hypoxia-, monocrotaline (MCT)-Sugen+hypoxia (SuHx)-*Gdf2-/-* and *Gdf2+/+* rats. Lower panels show quantification of cardiomyocyte cross-sectional area and picrosirius positive area. Data are represented as mean± SEM. Scale bar = 50µm in all sections. Significance was measured using 1-way ANOVA with Tukey post hoc tests. *, p<0.05; **, p<0.01; ***, p<0.001; ****, p<0.0001 *versus Gdf2+/+* control rats. $, p<0.05; $$, p<0.01; $$$, p<0.001; $$$$, p<0.0001 *versus Gdf2*+/+ CHx, MCT, or SuHx rats.

Because the vascular remodeling and PH induced by chronic hypoxia are known to be relatively mild and reversible, we next evaluated the susceptibility of *Gdf2*-/-rats to develop irreversible remodeling of the pulmonary vessels in two severe models of experimental PH induced by monocrotaline (MCT) and by a single injection of SU5416 before exposure to hypoxia for 3 weeks (SuHx). No rats died during these studies. Compared with vehicle-treated *Gdf2*-/- and *Gdf2*+/+ rats in room air, evidence of PH was found in *Gdf2*-/-as well as in *Gdf2*+/+ MCT and SuHx rats. This was reflected by increases in mPAP, TPVR and RV/(LV+S) ratio, together with a decrease in CO, without changes of systemic blood pressure. However, this elevation in mPAP and TPVR was less pronounced in *Gdf2*-/-MCT and SuHx rats (**Figure 6**). Consistent with these results, *Gdf2*-/-MCT and SuHx rats displayed lower percentages of wall thickness and of muscularized distal pulmonary arteries relative to *Gdf2*+/+ MCT and SuHx rats (**Figure 7A**). Furthermore, *Gdf2*-/-MCT and SuHx rats displayed less pronounced increase in the accumulation of collagen (stained with picroSirius red) in the RV **(Figure 7B)**.

*Gdf2*-/-rats are therefore less susceptible than wild-type rats to the elevation of pulmonary vascular resistance induced by either CHx, MCT, and SuHx.

## Discussion

A significant body of evidence supports the notion that weakening BMP-ALK1-Smad1/5/8 signaling in pulmonary microvascular endothelial cells (PMECs) contributes to various pathogenic features of pulmonary arterial hypertension (PAH), such as endothelial dysfunction, vascular smooth muscle cell proliferation, and resistance to apoptosis. Our study showed that BMP-9 deficiency in rats caused alterations in the 3D architecture of the lung microcirculation, leading to vasodilation and increased vascular density. We also confirmed that BMP-9 loss in rodents is not sufficient to cause spontaneous PH. Additionally, we extended previous studies in mice ^18, 19^ and demonstrated that *Gdf2* knockout rats exhibit a less severe increase in pulmonary pressures and vascular resistance compared to *Gdf2*+/+ rats when irreversible remodeling and severe PH are induced by MCT and SuHx. Furthermore, we showed that BMP receptor abundance, particularly ALK1, influenced both the endothelial transcriptional landscape and transcriptional responses to BMP-9 in human PMECs. Our observations also revealed that, similar to BMP-9 levels, the abundance of ALK1 had a significant impact on PMEC tube formation, migration, and proliferation. Moreover, our study highlighted a complex interplay existing between BMP-9, ALK1, and vascular endothelial growth factor (VEGF)/VEGFR activities.

ALK1-mediated endothelial BMP-9 signaling through Smad1/5/8 activation is well recognized to be critical for the maintenance of the endothelial integrity and stability, especially for the pulmonary endothelium that express high levels of BMPR-II, ALK1, and endoglin ^20^. Consistent with this notion, loss-of-function mutations in *BMPR2*, *ALK1*, *ENG*, *GDF2*, *BMP10* and *CAV1* are known predisposing factors for the structural and functional remodeling of the pulmonary circulation that are characteristic of PAH. Also, loss-of-function mutations in *ALK1*, *ENG, or SMAD4*, and more rarely in *GDF2*, are known causes of hereditary hemorrhagic telangiectasia (HHT)-a rare vascular multisystemic disease that leads to epistaxis, anemia due to blood loss, and arteriovenous malformations (AVMs) in organs such as the lungs, liver and brain ^21, 22^. Finally, recent reports of isolated pulmonary AVMs associated with *BMPR2* pathogenic variants highlight the complex pulmonary vascular consequences of genetic defects in the BMP–ALK1–Smad1/5/8 pathway ^23^. Although PAH and HHT share defects of key common members of the BMP–ALK1–Smad1/5/8 pathway, the reasons of their markedly different clinical presentation and penetrance (<30% in PAH and 100% in HHT) are still currently unknown. Similarly, the reasons why human findings are challenged by experimental evidence in rodents remain debated. In mice, the knockdown of BMP-9 alone or associated with the loss of its structural analogue BMP-10, is indeed not sufficient to lead to pulmonary vascular remodeling or the development of PH and unexpectedly protected against PH-induced by chronic hypoxia ^18, 19^. Here, we partly reconcile some of these human and mouse observations by providing *in vivo* evidence in *Sprague Dawley* rats that BMP-9 is a negative regulator of pulmonary vascular network formation and a factor that promotes pulmonary vasoconstriction. Even if *Gdf2*-/-rats developed and grew normally with no obvious defects in appearance, size, or fertility, a careful examination of the pulmonary arterial tree indeed showed higher vessel volume and numbers of vascular junctions when compared with *Gdf2*+/+ rats. This increased microvascular density and vasodilation may partly explain the less pronounced elevation in pulmonary vascular resistance and pressure in experimental PH in BMP-9 deficient rodents. These findings confirm and extend previous observations obtained in *Bmp-9* knockout mice and in C57BL/6 mice treated with neutralizing anti-BMP-9 antibodies showing the absence of spontaneous PH under normoxic conditions and the partial protection found in chronic hypoxia-induced PH model ^18, 19^. These studies have reported that BMP-9 deficiency or its inhibition were associated with lower levels of endothelin (ET)-1 and higher levels of two potent vasodilator factors, apelin and adrenomedullin (ADM) ^18, 19^. Here, we also showed that *Gdf2*-/-rats were partially protected against vascular remodeling-and PH-induced by chronic hypoxia, and extent these observations by showing that they were also partially protected against irreversible remodeling of the pulmonary vasculature and severe PH induced by MCT and SuHx.

The pulmonary microvascular endothelium is metabolically highly active, sensing and responding to signals from extracellular environments by secreting the correct substances by which it may maintain the vasomotor balance and vascular-tissue homeostasis ^24, 25^. Therefore, PMECs have heterogeneous phenotypes that match the diverse structures and metabolic needs of each vascular beds present in the lungs. Consistent with this notion, it was recently reported that primary cultures of human pulmonary artery endothelial cells (PAECs) isolated from the main pulmonary artery and 1^st^– 4^th^ branches exhibited 4 distinct transcriptomic profiles among 8 different PAEC-clusters ^26^. Here, primary cultures of PMECs derived from the distal parenchyma of human lungs (used at early passages < 4; n=5 subjects) in which small and medium-sized vessels are present were used. Our scRNA-seq analyses revealed 5 distinct PMEC-populations with 5 specific transcriptomic profiles. Because the response to BMP depends on which type-1, type-2 and type-3 receptors are expressed by the signal-receiving cell, we used the PMEC diversity present in primary cultures of human cells to analyze the BMP-9 transcriptomic differences between ALK1^high^ and ALK1^low^ PMECs. ALK1 was indeed found to be the predominant type-1 receptor expressed, along with BMPR-II, TGFβRII and endoglin and its abundance was ∼1.5 higher in the cluster-5 (ALK1^high^ PMECs) when compared with the other PMEC-clusters (ALK1^low^ PMECs). A careful examination of the cellular phenotype in ALK1^high^ PMECs indicated that C5 exhibited a pro-angiogenic signature at baseline that was attenuated in presence of exogenous BMP-9. A substantial response to BMP-9 was also observed in ALK1^low^ PMECs with specific transcriptomic signatures between the 4 distinct PMEC-clusters. Consistent with the notion that ALK1 is known to be a high-capacity receptor for low-density lipoprotein (LDL) ^27, 28^, several GO/BP terms related to fatty acid and lipid transport and homeostasis were found in ALK1^low^, but not in ALK1^low^ PMECs treated with BMP-9. Indeed, BMP-9 can control cell surface levels of ALK-1, via caveolin-1 to regulate both BMP-9 signaling and LDL transcytosis ^27^. The effect of BMP-9 on ALK1 levels might partly explain why the pulmonary endothelium of *Gdf2*-/-rats exhibited higher ALK1 levels than *Gdf2*+/+ rats. As already reported ^17^, we also found that BMP-9 participated in a positive feedback loop that increases BMPR-II in PMECs. Here, the same was noted for endoglin. These BMP-9-mediated changes in BMP receptors point out that the BMP-9 signaling is dynamically controlled by complex regulatory mechanisms. Using functional *in vitro* studies, we validated some of these scRNA-seq findings. Consistent with previous studies ^7^, we first found that addition of exogenous BMP-9 on human PMEC cultures attenuated cell migration, proliferation and tube formation, while its removal with ALK1-Fc showed the opposite effects. Second, we found that attenuation of ALK1 acts as a critical modulator of endothelial phenotype, as reflected by the reduced PMEC migration, proliferation and tube formation observed with ALK1 siRNA-mediated knockdown or inhibition with ML 347. In the same line, a recent study has demonstrated that BMP-9 could act as an angiogenic switch that can either promote or prevent EC proliferation, depending of the BMPR-II level ^29^. High levels of BMPR-II and/or ALK1 at the EC surface are therefore two important factors associated with high angiogenic potential of PMECs, which signaling is finely regulated by BMP-9.

Regulation of vascular development and pathological angiogenesis are regulated by many angiogenic factors, among which signals induced by the VEGF and BMP-9/ALK1 appear to play an important role. Consistent with the fact that enhanced VEGF signaling has already been associated with formation of large, dilated, and fragile blood vessels, which resembled AVMs ^30–32^, we found that *Gdf2-/-* rat lungs displayed higher VEGF/VEGFR activities relative to *Gdf2*+/+. In addition, we found that exogenous BMP-9 was able to reduce the phosphorylation of VEGFR2 and its downstream signaling through SRC and AKT in cultured human PMECs. We also found that the ALK1^high^ PMEC-cluster expressed higher levels of several VEGF ligands including of VEGF-C and placental growth factor (PGF), but also of VEGFR1 and VEGFR2 than the ALK1^low^ PMEC-cluster. Interestingly, we also found that BMP-9 inhibited levels of VEGFR2, p-SRC, and p-AKT in human PMECs exposed to VEGF. The fact that BMP-9 and ALK1 are two strong modulators of VEGF-C expression is consistent with the fact that *Bmp9* KO mice have been reported to exhibit defects in lymphatic capillaries and collecting vessel maturation as well as in valve formation ^33–36^. These observations are consistent with the key role played by a crosstalk between BMP-9/ALK1 and VEGF systems in HHT and other vascular diseases. Other studies have indeed also highlighted the intricate involvement of BMP-9/ALK1 in stabilizing the arteriovenous network, with this effect being dependent on the specific vascular bed^37, 38^. However, these crosstalks between BMP-9/ALK1 and VEGF systems can also contribute to the loss of the small pulmonary vessels in experimental and human PAH, through a mechanism entitled vascular “pruning” that leads to a pattern of vascular rarefaction (image in “dead tree”), a phenomenon that further exacerbates the elevation of pulmonary vascular resistance. In PAH, it is indeed suspected that the uncontrolled and chronic activation of the VEGF, and other pro-angiogenic pathways, might have deleterious effects on the formation of new vessels *per se* ^39–41^. VEGF is indeed abundant in the lung, and a known critical factor for the maintenance of the pulmonary structures. A recent study has underlined that loss-of-function variants in *KDR* (encoding VEGFR2) are associated with a particular form of PAH characterized by a range of lung parenchymal abnormalities, including small airways disease, emphysema and mild pulmonary fibrosis ^42^. A better understanding on how specific environmental and pathological factors present in HHT and PAH may influence the crosstalk between BMP-9/ALK1 and VEGF systems and therefore the disease onset, progression, and manifestation is clearly needed.

BMP-9 has also other functions, such as the ability to prevent vascular permeability, to attenuate EC apoptosis, and to maitain fenestration of sinusoidal liver ECs ^43^. It also has been shown to causes the increase of vascular permeability in the lung ^17^, and to protect ECs against the deleterious effect of inflammation by decreasing certain chemoattractants of leukocytes such as C-C motif chemokine ligand 2 (CCL2) ^44^. Thus, beyond the scope of this study, there is a need for research to examine more in details the specific roles and actions of BMP-9 in the preservation of other tissue, organ, structure integrity.

Taken together, our data identify BMP-9 and ALK1 as two critical factors for pulmonary vascular growth and remodeling *in vivo*. Our study is the first to point out that the higher pulmonary vessel density and vasodilation could partly explained why animals deficient in BMP-9 display lower susceptibility to develop pulmonary vascular remodeling and experimental PH. We also provided *in vitro* and *in vivo* molecular evidence that this phenotype is driven by a higher ALK1 expression and activation of the VEGFR2 signaling. This intricate interplay between the BMP/TGFβ and VEGF/VEGR pathways offers new possibilities for developing PAH medications targeting the TGF-β superfamily, with the aim restoring pulmonary vascular homeostasis ^45–47^.

## Data Availability

The data utilized in this study is accessible upon request. In light of privacy and confidentiality concerns, we are unable to openly distribute the dataset. However, researchers interested in accessing the data can reach out to the corresponding author for requesting access. Each request will be thoroughly reviewed, and we will make every effort to provide the required data while adhering to relevant regulations and ethical considerations.

## Acknowledgments

The authors thank Valérie Domergue and her team from the animal facility of UMS IPSIT for their help with the animals. The authors acknowledge Vincent Thomas de Montpreville and all pathologists and technicians from the Centre de Recherche Biologique at Marie Lannelongue Hospital - Groupe Hospitalier Paris Saint Joseph for their expertise and support. They also thank the French PAH patient association (HTAP France) and all participants of the French PH Network PulmoTension.

## Authors’ contributions

Conception and design: NB, LT, CG; Analysis and interpretation: all; Drafting manuscript: NB, LT, CG. SR and IA generated the *Gdf2*-/-KO rats.

## Supplemental Materials

*Table S1-2* Figure S1-6

